# An electrocardiography score predicts heart failure hospitalization or death beyond that of cardiovascular magnetic resonance imaging

**DOI:** 10.1101/2022.01.04.22268654

**Authors:** Maren Maanja, Todd T Schlegel, Fredrika Fröjdh, Louise Niklasson, Björn Wieslander, Ljuba Bacharova, Erik B Schelbert, Martin Ugander

**Affiliations:** Department of Clinical Physiology, Karolinska University Hospital, and Karolinska Institutet, Stockholm, Sweden; Nicollier-Schlegel SARL, Trélex, Switzerland; International Laser Center, and Institute of Pathophysiology, Comenius University, Bratislava, Slovak Republic; Department of Medicine, University of Pittsburgh Medical Center, Pittsburgh, PA, USA; The Kolling Institute, Royal North Shore Hospital, and Northern Clinical School, Sydney Medical School, University of Sydney, Sydney, Australia

**Keywords:** Electrocardiography (ECG), Magnetic Resonance Imaging (MRI), Prognosis

## Abstract

**Background:** The electrocardiogram (ECG) and cardiovascular magnetic resonance imaging (CMR) both provide powerful prognostic information. The aim was to determine the relative prognostic value of ECG and CMR, respectively.

**Methods:** Consecutive patients (n=783) undergoing CMR and resting 12-lead ECG with a QRS duration <120 ms were included. CMR measures included feature tracking global longitudinal strain (GLS), extracellular volume fraction (ECV), left ventricular mass and volumes, and ischemic and non-ischemic scar size. Prognosis scores for one-year event-free survival were derived using continuous ECG or CMR measures, and multinomial logistic regression, and compared with regards to the combined outcome of survival free from hospitalization for heart failure or death.

**Results:** Patients (median [interquartile range] age 55 [43–64] years, 44% female) had 155 events during 5.7 [4.4–6.6] years. The ECG prognosis score included 1) the frontal plane QRS-T angle, and 2) the heart rate corrected QT duration (QTc) (log-rank 55, *p*<0.001). The CMR prognosis score included 1) GLS, and 2) ECV (log-rank 85, *p*<0.001). The combination of positive scores for both ECG and CMR yielded the highest prognostic value (log-rank 105, *p*<0.001). Multivariable analysis showed an association with outcomes for both the ECG prognosis score (log-rank 8.4, hazard ratio [95% confidence interval] 1.29 [1.09–1.54], *p*=0.004) and the CMR prognosis score (log-rank 47, hazard ratio 1.90 [1.58–2.28], *p*<0.001).

**Conclusions:** An ECG prognosis score predicted outcomes independently of, and beyond CMR. Combining the results of ECG and CMR using both prognosis scores improved the overall prognostic performance.

## Introduction

Prognostic information can be gleaned from both the electrocardiogram (ECG) (1) and cardiovascular magnetic resonance imaging (CMR) measures of left ventricular mass (2), cardiac function (3), and myocardial tissue characterization (4). However, the ECG and CMR provide potentially complementary information, and how these measures perform in relation to each other has not been studied head-to-head.

The ECG reflects the electrical activity of the heart, and the information present in the ECG may complement measures of cardiac structure and function. To study the prognostic performance, prognosis scores can be created from parameters that best predict outcome. We aimed to create separate prognosis scores for the ECG and CMR, respectively, and compare them with regards to ability to predict the combined end-point of hospitalization for heart failure or death. Furthermore, rather than generating scores based on attributing points to criteria fulfilling set thresholds (point-based scores), we sought to harness the statistical strength of continuous variables to create continuous scores ranging from 0–100% likelihood of an event. We also sought to compare our results to a recently published point-based ECG risk score (5). The hypothesis of the study was that an ECG score could provide incremental prognostic information beyond CMR measures with known powerful prognostic performance.

## Methods

### Study patients

In this cross-sectional study, patients were identified from a prospectively acquired database of 1828 enrolled patients referred for a clinical CMR scan at University of Pittsburgh Medical Center (UPMC; Pittsburgh, PA, USA) between 2008 and 2017, and followed until April 2018. The study was approved by the UPMC Institutional Review Board, and all participants provided written informed consent. Inclusion criteria were completion of a gadolinium-based contrast agent-enhanced CMR, and a digital ECG with sinus rhythm and a QRS duration <120 ms acquired within 30 days of CMR. Exclusion criteria were ECG confounders such as atrial fibrillation or flutter, paced rhythm, severe arrhythmia defined as premature atrial or ventricular contractions in bigeminy or trigeminy. Further exclusion criteria were CMR findings of amyloidosis, hypertrophic cardiomyopathy, Takotsubo cardiomyopathy, congenital heart disease, siderosis, Fabry’s disease and poor CMR image quality. Hospitalization for first heart failure event after CMR was identified by medical record review using pre-defined criteria evaluated by a board-certified cardiologist, and mortality status was ascertained by medical record review and Social Security Death Index queries as previously described (6).

### Subgroups

From the database, 783 patients had an ECG within 30 days of CMR with a sinus rhythm and QRS <120 ms and were included to create the ECG score. Among these, 730 patients had readily available CMR data including global longitudinal strain (GLS) and extracellular volume fraction (ECV), and were used to create the CMR score, and for prognosis calculations. For the ECG and CMR prognosis scores, respectively, patients were grouped as either having experienced an event of hospitalization for heart failure or death within one year of CMR, or no event.

### CMR acquisition and analysis

CMR imaging methodology and typical acquisition parameters have previously been described (7). In brief, CMR images were acquired using a 1.5 Tesla scanner, and LV mass, volumes, and ejection fraction were measured from short-axis stacks of end-systolic and end-diastolic cine images. ECV was calculated in basal and mid-ventricular myocardial short-axis slices in areas without focal late gadolinium enhancement (LGE), as described previously (8). GLS analysis was performed using semi-automated tissue feature tracking software in end-diastole. For an extended description of CMR methodology, see the supplemental material.

The following CMR parameters were imputed into the CMR prognosis score analysis: GLS, LVEF, ECV, left ventricular end-diastolic volume indexed to body surface area (LVEDVI), left ventricular mass indexed to body surface area (LVMI), infarct size by LGE, and non-ischemic scar size by LGE.

### ECG acquisition and analysis

Resting 12-lead ECG data for each subject was collected from the local ECG storage system (MUSE® Cardiology Information System, Version 8.0 SP2, GE Healthcare, IL, USA) and exported into anonymized xml files with coded identification. The xml files were analyzed digitally using semi-automated software developed in-house. The following ECG measures were included into the ECG prognosis score analysis: conventional ECG parameters, vectorcardiographic measures derived using Kors’ transform (9), and QRS-wave and T-wave complexity measures quantified using singular value decomposition (10). The combination of the aforementioned ECG methods is referred to as Advanced-ECG (A-ECG). The conventional ECG risk score by Aro et al (5) (Aro ECG risk score), was also investigated, and defined as fulfillment of at least four of the following: heart rate >75 bpm, QRS duration >110 ms, QTc time > 450 ms in men and > 460 in women, Tpeak–Tend duration >89 ms, frontal plane QRS-T angle >90°, delayed QRS transition zone, electrocardiographic left ventricular hypertrophy (LVH), or a delayed intrinsicoid deflection.

#### Statistical analysis

Statistical analysis was performed in R (R Foundation for Statistical Computing, version 3.4.3, Vienna, Austria). Prognosis scores for one-year event-free survival were derived using multinomial stepwise forward logistic regression for variable selection. The event time of one year was chosen for investigation as it is at a time point when both a statistically and clinically meaningful number of events would have been observed. The area under the receiver operating curve (AUC) was bootstrapped 2000 times to obtain the 95% confidence intervals (CI). In order to avoid overfitting the score, one incremental parameter was considered appropriate for every ten events (11). The ECG or CMR parameters, respectively, that yielded the highest AUC were chosen for the respective prognosis scores. The Youden index (12) or, when not applicable, the point closest the top left in the AUC (13), was used to determine the score cut-off that optimizes sensitivity and specificity for a given score. DeLong’s test was used to compare the AUC of ECG prognosis score to the Aro ECG risk score. The prognosis scores ranged from 0% to 100% and showed the likelihood of having an event as mathematically calculated from the logistic regression by:

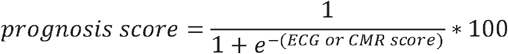

The constants in the logistic regression equations for both the ECG and CMR score were adjusted so that the optimal cut-offs for event risk were defined as ≥50%. The prognosis scores were compared to each other with regards to survival free from hospitalization for heart failure or death using univariable and multivariable Cox regression, and Kaplan-Meier analysis. The chi-square (*χ*^2^) value was calculated to compare the effect of the estimates. Hazard ratios were reported with an increment of one standard deviation (SD) of the respective measures. Linear correlations were evaluated using Pearson’s correlation coefficient and expressed as its square (R^2^). Multivariable linear regression was performed to investigate how CMR measures related to the ECG prognosis score. The Kolmogorov-Smirnov test was used to test if data were normally distributed, and differences between subgroups’ baseline data were tested using the Mann-Whitney U test or chi-square test, as appropriate. Data were described using median and interquartile range, or percentage, respectively. Furthermore, prediction models using logistic regression with a Ridge, Lasso, or Elastic Net penalty, respectively, as well as Cox regression with a Lasso penalty were also evaluated. A *p*-value <0.05 was considered statistically significant.

## Results

Baseline characteristics of the study population are presented in Table 1. A total of 783 patients with a CMR scan and an ECG with a QRS duration <120 ms and without rhythm confounders were included in the study. All 783 patients were used to create the ECG score, and 730 patients were used to create the CMR score and for the survival analyses. The study patients experienced 155 events during 5.7 [4.4–6.6] years follow-up: 113 deaths (14.4%), 68 (8.7%) hospitalizations for heart failure, and 26 (3.3%) with both. In comparison to patients with no event, patients with an event were older, and had more CMR comorbidities and cardiovascular medications.

**Table 1.**
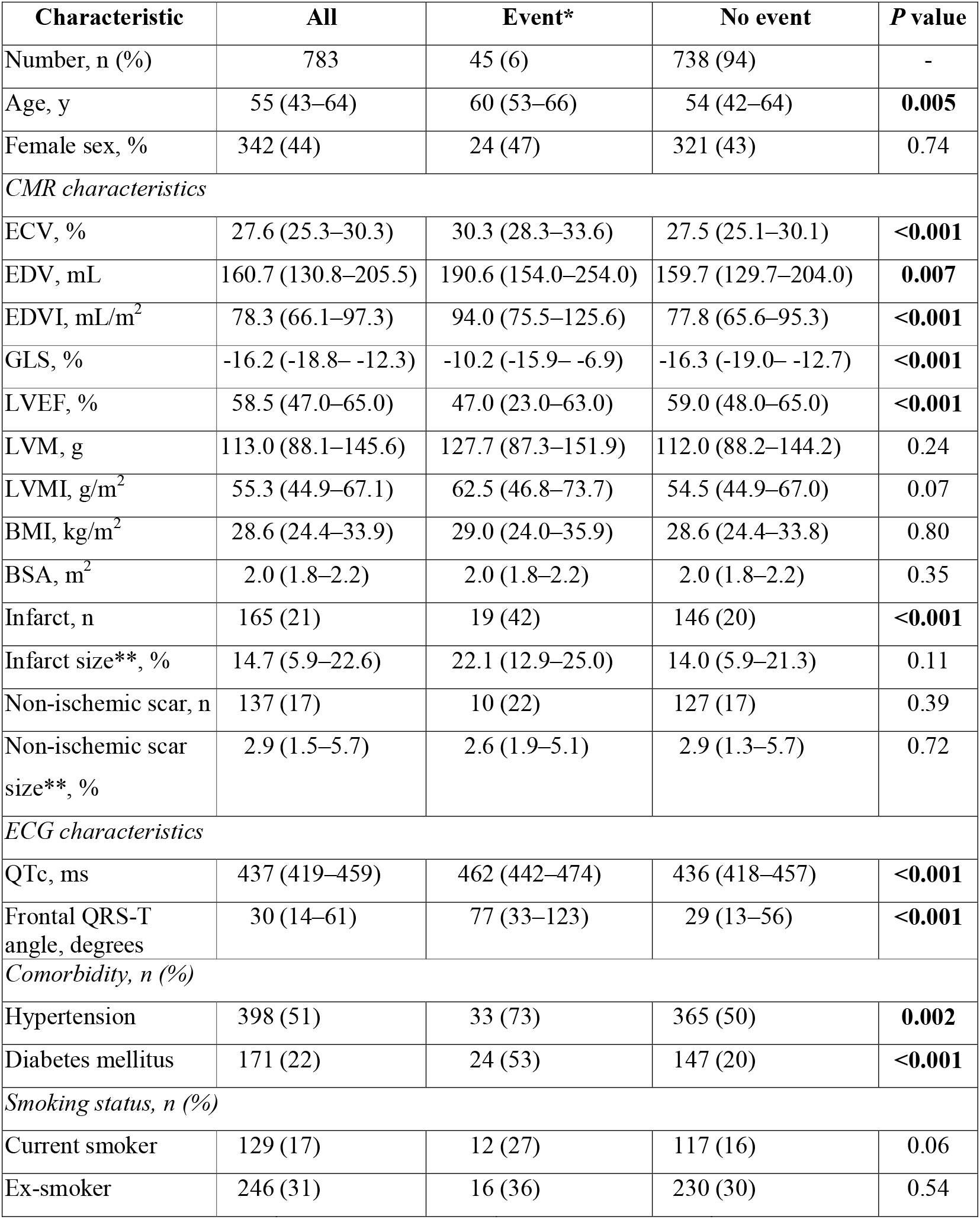
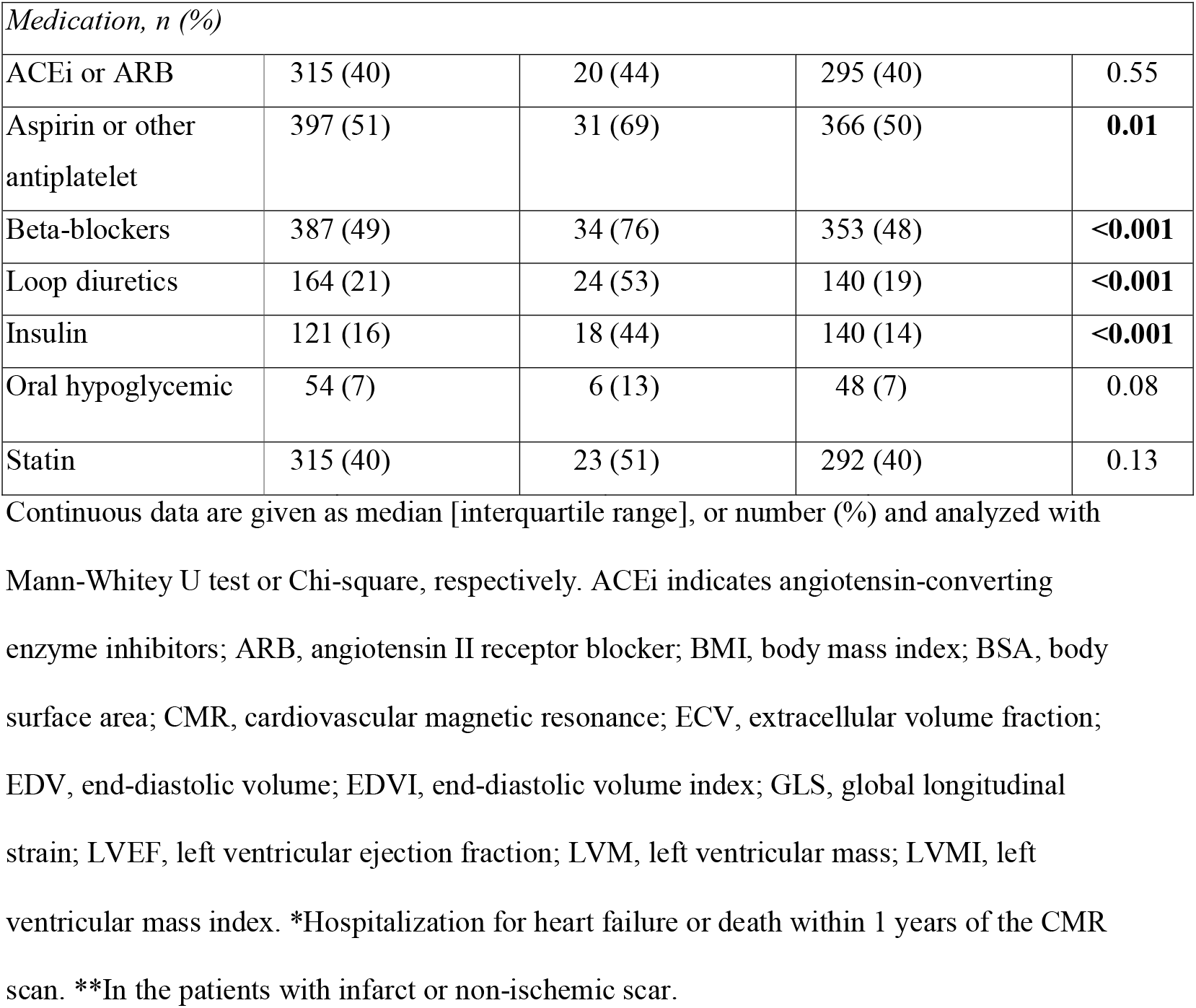
Baseline characteristics for the study population.

| Characteristic | All | Event* | No event | P value |
| --- | --- | --- | --- | --- |
| Number, n (%) | 783 | 45 (6) | 738 (94) | - |
| Age, y | 55 (43–64) | 60 (53–66) | 54 (42–64) | **0.005** |
| Female sex, % | 342 (44) | 24 (47) | 321 (43) | 0.74 |
| *CMR characteristics* |  |  |  |  |
| ECV, % | 27.6 (25.3–30.3) | 30.3 (28.3–33.6) | 27.5 (25.1–30.1) | **<0.001** |
| EDV, mL | 160.7 (130.8–205.5) | 190.6 (154.0–254.0) | 159.7 (129.7–204.0) | **0.007** |
| EDVI, mL/m<sup>2</sup> | 78.3 (66.1–97.3) | 94.0 (75.5–125.6) | 77.8 (65.6–95.3) | **<0.001** |
| GLS, % | -16.2 (-18.8– -12.3) | -10.2 (-15.9– -6.9) | -16.3 (-19.0– -12.7) | **<0.001** |
| LVEF, % | 58.5 (47.0–65.0) | 47.0 (23.0–63.0) | 59.0 (48.0–65.0) | **<0.001** |
| LVM, g | 113.0 (88.1–145.6) | 127.7 (87.3–151.9) | 112.0 (88.2–144.2) | 0.24 |
| LVMI, g/m<sup>2</sup> | 55.3 (44.9–67.1) | 62.5 (46.8–73.7) | 54.5 (44.9–67.0) | 0.07 |
| BMI, kg/m<sup>2</sup> | 28.6 (24.4–33.9) | 29.0 (24.0–35.9) | 28.6 (24.4–33.8) | 0.80 |
| BSA, m<sup>2</sup> | 2.0 (1.8–2.2) | 2.0 (1.8–2.2) | 2.0 (1.8–2.2) | 0.35 |
| Infarct, n | 165 (21) | 19 (42) | 146 (20) | **<0.001** |
| Infarct size**, % | 14.7 (5.9–22.6) | 22.1 (12.9–25.0) | 14.0 (5.9–21.3) | 0.11 |
| Non-ischemic scar, n | 137 (17) | 10 (22) | 127 (17) | 0.39 |
| Non-ischemic scar size**, % | 2.9 (1.5–5.7) | 2.6 (1.9–5.1) | 2.9 (1.3–5.7) | 0.72 |
| *ECG characteristics* |  |  |  |  |
| QTc, ms | 437 (419–459) | 462 (442–474) | 436 (418–457) | **<0.001** |
| Frontal QRS-T angle, degrees | 30 (14–61) | 77 (33–123) | 29 (13–56) | **<0.001** |
| *Comorbidity, n (%)* |  |  |  |  |
| Hypertension | 398 (51) | 33 (73) | 365 (50) | **0.002** |
| Diabetes mellitus | 171 (22) | 24 (53) | 147 (20) | **<0.001** |
| *Smoking status, n (%)* |  |  |  |  |
| Current smoker | 129 (17) | 12 (27) | 117 (16) | 0.06 |
| Ex-smoker | 246 (31) | 16 (36) | 230 (30) | 0.54 |

| *Medication, n (%)* |  |  |  |  |
| --- | --- | --- | --- | --- |
| ACEi or ARB | 315 (40) | 20 (44) | 295 (40) | 0.55 |
| Aspirin or other antiplatelet | 397 (51) | 31 (69) | 366 (50) | **0.01** |
| Beta-blockers | 387 (49) | 34 (76) | 353 (48) | **<0.001** |
| Loop diuretics | 164 (21) | 24 (53) | 140 (19) | **<0.001** |
| Insulin | 121 (16) | 18 (44) | 140 (14) | **<0.001** |
| Oral hypoglycemic | 54 (7) | 6 (13) | 48 (7) | 0.08 |
| Statin | 315 (40) | 23 (51) | 292 (40) | 0.13 |

Continuous data are given as median [interquartile range], or number (%) and analyzed with
Mann-Whitey U test or Chi-square, respectively. ACEi indicates angiotensin-converting enzyme inhibitors; ARB, angiotensin II receptor blocker; BMI, body mass index; BSA, body surface area; CMR, cardiovascular magnetic resonance; ECV, extracellular volume fraction; EDV, end-diastolic volume; EDVI, end-diastolic volume index; GLS, global longitudinal strain; LVEF, left ventricular ejection fraction; LVM, left ventricular mass; LVMI, left ventricular mass index. \*Hospitalization for heart failure or death within 1 years of the CMR scan. \*\*In the patients with infarct or non-ischemic scar.

### ECG prognosis score

The final ECG prognosis score for 1-year event included 1) the frontal plane QRS-T angle (degrees), and 2) the heart rate corrected (Bazett) QT duration (ms), and was calculated as:

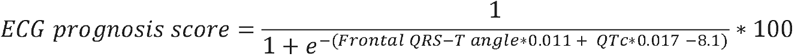

The ECG prognosis score had an AUC (95% CI) of 0.78 (0.71–0.84), see Figure 1, a sensitivity of 78% (67–89%) and a specificity of 71% (62–80%). The Frontal QRS-T angle and QTc were negligibly correlated (R^2^=0.06, *p*<0.001). In multivariable linear regression analysis, the ECG prognosis score related to GLS, ECV, LVMI and infarct size (global R^2^=0.28, *p*<0.001). An additional ECG score was also created with four ECG parameters, which was the maximum number that did not violate the rule of 10 events per variable. However, the more complex four-parameter ECG score did not add a meaningful magnitude of incremental prognostic power (results not shown).

**Figure 1.**
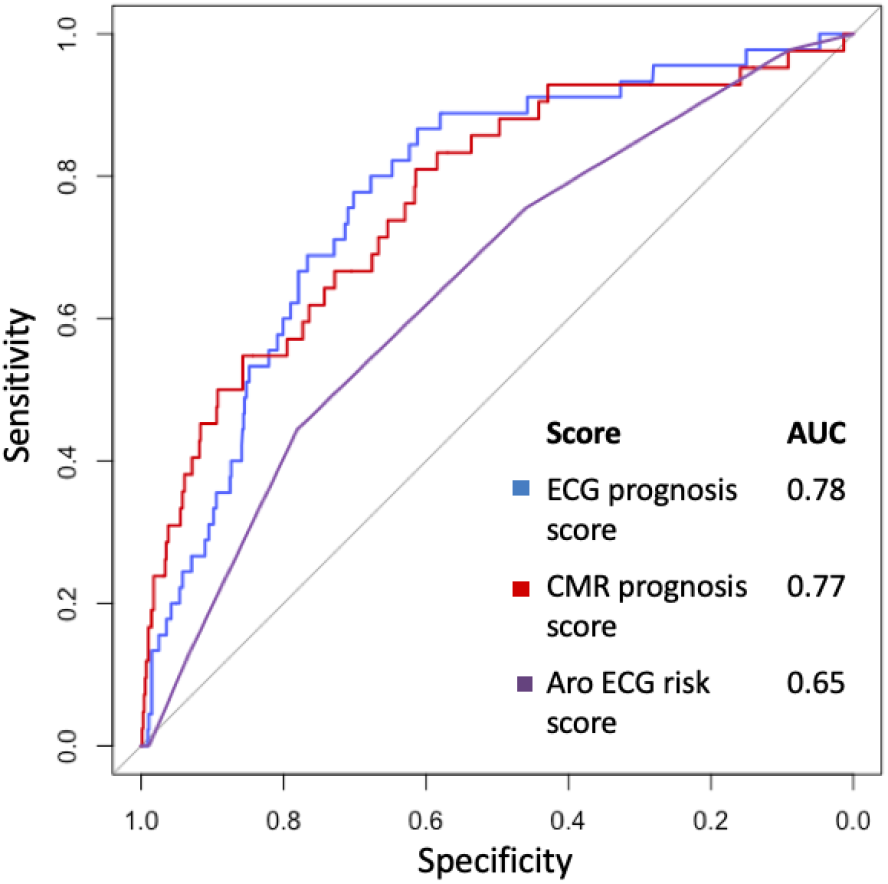
Receiver operating characteristics curve for the ECG and CMR prognosis scores, respectively, and the Aro ECG risk score. The area under the curve (AUC) for the ECG prognosis score was greater than for the Aro ECG risk score, p<0.001.

An A-ECG score based on Cox regression included a combination of 19 parameters, and had an AUC (95% CI) of 0.77 (0.69 – 0.84) after one year of follow-up. The prediction models using logistic regression with a Ridge, Lasso, or Elastic Net penalty, respectively, did either not outperform multinomial stepwise forward logistic regression, or did not provide any meaningful magnitude of improved diagnostic performance (results not shown).

### CMR prognosis score

The final CMR prognosis score for 1-year event included 1) GLS (%), and 2) ECV (%), and was calculated as:

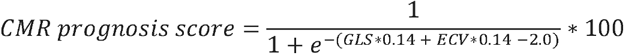

The CMR prognosis score had an AUC of 0.77 (0.69–0.84), see Figure 1, a sensitivity of 79% (50–95%) and a specificity of 65% (46–92%). GLS and ECV were negligibly correlated (R^2^=0.05, *p*<0.001).

### Aro ECG risk score

Fifty-two patients (7%) out of the total study population, and 19 patients (12%) with an event had four or more abnormal parameters in the Aro ECG risk score. The Aro ECG risk score yielded an AUC of 0.65 (0.57–0.72), see Figure 1, a sensitivity of 12% and a specificity of 93%. Univariable Cox showed an association with outcomes (χ^2^ 14, HR 1.33 [1.14–1.55], *p*<0.001). In the current study, the Aro ECG risk score had a lower AUC than the ECG prognosis score, *p*<0.001.

### ECG and CMR prognosis scores and outcomes

Table 2 summarizes the results for the Cox regressions. Univariable Cox analysis showed that the CMR prognosis score had a higher univariable association with outcomes (χ^2^ 93, HR 2.17 [1.85–2.54], *p*<0.001) than the ECG prognosis score (χ^2^ 58, HR 1.79 [1.54–2.08], *p*<0.001). Multivariable Cox regression showed an association with outcomes for both the CMR prognosis score (χ^2^ 47, *p*<0.001) and the ECG prognosis score (χ^2^ 8.4, *p*=0.004), respectively. Kaplan Meier analysis showed that the combination of a score ≥50% for both the ECG (log-rank 55, *p*<0.001) and the CMR prognosis score (log-rank 85, *p*<0.001) yielded the highest prognostic value (log-rank 105, *p*<0.001). For the event of hospitalization for heart failure only, or death only, respectively, the ECG prognosis score had a log-rank of 43, and log-rank of 31, respectively, *p*<0.001 for both.

**Table 2.** Univariable and multivariable Cox regression for the ECG and CMR prognosis scores, as well as their respective components. The HR increment was 1 SD of the respective measure.

|  |  | Univariable Cox regression |  |  | Multivariable Cox regression |  |  |
| --- | --- | --- | --- | --- | --- | --- | --- |
| Variable | Increment for HR | $\chi^2$ | HR (95% CI) | *p*-value | $\chi^2$ | HR (95% CI) | *p*-value |
| **ECG prognosis score** | 0.18 | 58 | 1.79 (1.54-2.08) | <0.001 | 8.4 | 1.29 (1.09-1.54) | 0.004 |
| Frontal QRS-T angle | 44 | 38 | 1.48 (1.30-1.69) | <0.001 | - | - | - |
| QTc | 32 | 36 | 1.60 (1.37-1.88) | <0.001 | - | - | - |
| **CMR prognosis score** | 0.20 | 93 | 2.17 (1.85-2.54) | <0.001 | 47 | 1.90 (1.58-2.28) | <0.001 |
| GLS | 4.7 | 73 | 1.93 (1.66-2.24) | <0.001 | - | - | - |
| ECV | 3.9 | 46 | 1.66 (1.43-1.99) | <0.001 | - | - | - |

Figure 2 shows survival for the ECG prognosis score, the CMR prognosis score, and for patients with either a normal ECG and CMR score, an increased ECG or an increased CMR score, or both an increased ECG and CMR score, respectively, using score probability greater than 50% as the cut-off. Figure 3 shows survival for the Aro ECG risk score (log-rank 15, *p*=0.005), and the ECG (log-rank 64, *p*<0.001) and CMR (log-rank 103, *p*<0.001) prognosis scores, respectively, using score cut-offs at 25%, 50% and 75%.

**Figure 2.**
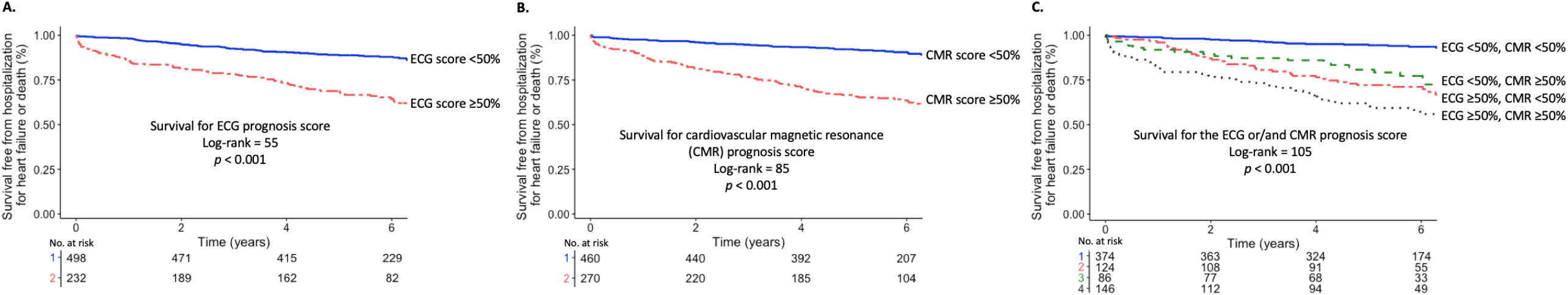
Kaplan Meier curves showing survival for **A**. the ECG prognosis score, **B**. cardiovascular magnetic resonance (CMR) prognosis score, and **C**. patients with normal ECG and CMR score, patients with either an increased ECG or an increased CMR score, and patients with both an increased ECG and CMR score.

**Figure 3.**
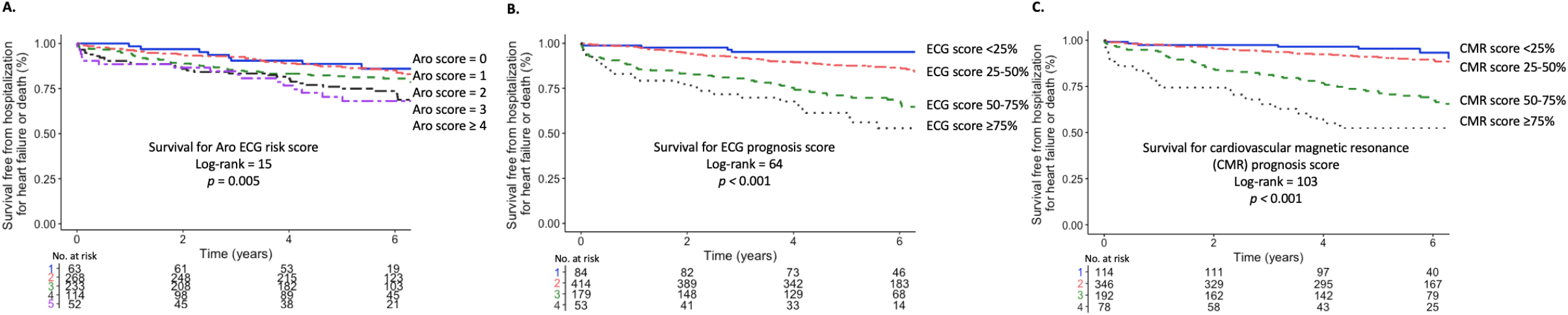
Kaplan Meier curves showing survival for **A**. the Aro ECG risk score, **B**. ECG prognosis score, and **C**. cardiovascular magnetic resonance (CMR) prognosis score.

## Discussion

The major finding of this study was that, in patients with a QRS duration <120 ms, a new ECG prognosis score provided prognostic information beyond CMR measures with known powerful prognostic utility. Importantly, the combination of the ECG prognosis score and the CMR prognosis score provided the best risk stratification, and should preferentially be used in concert.

### ECG measures and outcomes

The frontal plane QRS-T angle is the angle between the QRS axis and the T axis in the frontal plane. In a healthy heart, the angle between the QRS and T axis is small, whereas it increases as a result of myocardial pathology (14), reflecting an abnormal dispersion between the depolarization and repolarization of the left ventricle (15). Increased QRS-T angles are typically found in LVH, myocardial pacing, bundle branch blocks, and ischemia, and is a predictor of adverse events in patients with non-ischemic cardiomyopathy (16). The QT duration spans the beginning of the depolarization until the end of the repolarization. A prolonged heart rate corrected QT time, QTc (17), is associated with an increased risk of ventricular fibrillation and sudden death (18).

The ECG holds more proven prognostic information than is usually considered, e.g. in the vectorcardiographic spatial QRS-T angle (19, 20), and more complex measures of the QRS-wave and T-wave (21). However, the two-parameter ECG prognosis score from the current study did not include any such measure. It may be that such measures are associated with events over durations of time markedly shorter or longer than one year, since they are known to be associated with other specific cardiac diagnoses such as left ventricular hypertrophy, coronary artery disease, and left ventricular systolic dysfunction (10).

The Aro ECG risk score was constructed to predict sudden cardiac death, which may be why it had limited power in predicting the outcomes evaluated in the current study. Nevertheless, in the study by Aro, et al, 16% of all cases compared to 12% of the cases in the current study had four or more abnormal parameters. Furthermore, 3% of Aro et al’s controls compared to 7% of the patients without events in the current study had a risk score of four or more abnormal parameters (5). The control participants in the Aro, et al, study were individuals with no history of ventricular arrhythmias or cardiac arrest, whereas patients with no events in the current study where referred for a clinical CMR and are thus more likely to have cardiovascular disease. Besides, they may have had events prior to the cardiac scan (not investigated).

### CMR measures and outcomes

Cardiovascular disease can lead to an increase in the myocardial extracellular space, consisting mainly of diffuse myocardial fibrosis, leading to systolic and diastolic dysfunction, and an increased risk of arrhythmia and mortality (22). The prognostic utility of GLS has been proven to be superior to that of LVEF (23). GLS can detect changes in myocardial contractility that may proceed an abnormal LVEF (24). This might help explain why LVEF did not have a multivariable association with the CMR prognosis score in the presence of GLS, even though there was a difference in LVEF between patients with versus without a one-year event. Previous studies have reported LVMI as an outcome predictor (2), however, no difference in LVMI between patients with versus without one-year events was found in our study.

### Prognosis scores

Clinical cardiovascular risk scores are often based on multivariable algorithms of parameters readily available in clinics, such as age, blood pressure, and presence of diabetes (25). We used stepwise logistic regression to construct scores that estimate risk probabilities based on continuous rather than dichotomous variables. The benefits of doing this include reduced training and utilization times, and facilitation of data understanding (26). Developing a score based on Cox regression using a continuous outcome variable did not improve the performance of the score, and thus the logistic regression score was used in the interest of having a parsimonious score. Both measures incorporated into our ECG score are also components of the Aro ECG risk score. However, in spite of the greater complexity of the Aro ECG risk score, with incorporation of multiple additional dichotomous measures, its AUC was 0.65 in this study, compared to 0.78 for the simpler ECG prognosis score.

### ECG vs imaging for prognosis

The ECG has previously been compared to other imaging modalities with regards to prognosis, and found to have an incremental or independent prognostic power in, for instance, the setting of myocardial infarction with non-obstructed coronary arteries (27), and LVH (28). As the ECG and imaging provide different and complementary information, it is beneficial to assess both when possible.

### Study limitations

This study had several limitations. The retrospective design introduces the risk of selection bias, even though the data were acquired prospectively. This is a single-center study that may not reflect the general population, and patients with various ECG conditions such as bundle branch blocks, and atrial fibrillation were excluded due to a known adverse prognosis related to the arrhythmia as such (29). Furthermore, the risk scores were derived and validated in the same cohort. To account for this, the results were bootstrapped to obtain confidence intervals, although further validation in full separate cohorts is necessary. Nevertheless, all parameters incorporated into the scores have had proven prognostic utility in previous studies.

We also investigated the composite outcome of hospitalization for heart failure and all-cause mortality, which may not necessarily be due to cardiac pathology. However, all hospitalization events were adjudicated according to strict criteria.

### Clinical perspectives

The ECG and the parameters in the newly proposed ECG prognosis score, are readily presented numerically by most ECG vendors, making the ECG prognosis score a clinically accessible tool. Patients with a high score may benefit from more intensive risk management and surveillance, and future prospective studies are justified to evaluate such an approach. The ECG is less expensive and more available than CMR. However, the ECG and CMR convey independent and complementary prognostic information and should preferentially be combined when possible.

### Conclusions

A new ECG prognosis score predicted outcomes independently of, and beyond, comprehensive CMR measures with known prognostic power. Combining the results of ECG and CMR using both prognosis scores improved the overall prognostic utility.

## Supporting information

Detailed description of CMR methodology

## Data Availability

Data can be made available upon reasonable request.

## Nonstandard Abbreviations and Acronyms

A-ECG: Advanced Electrocardiography
CMR: Cardiovascular Magnetic Resonance
ECV: Extracellular Volume fraction
GLS: Global Longitudinal Strain
LGE: Late Gadolinium Enhancement
LV: Left Ventricle

## Acknowledgements

The authors acknowledge Daniel Loewenstein, MD, and Zak Loring, MD, for valuable discussions regarding the statistical analysis.

## References

1. Aro AL, Huikuri HV. Electrocardiographic predictors of sudden cardiac death from a large Finnish general population cohort. J Electrocardiol. 2013;46(5):434–8.

2. Armstrong AC, Gidding S, Gjesdal O, Wu C, Bluemke DA, Lima JA. LV mass assessed by echocardiography and CMR, cardiovascular outcomes, and medical practice. JACC Cardiovasc Imaging. 2012;5(8):837–48.

3. Kalam K, Otahal P, Marwick TH. Prognostic implications of global LV dysfunction: a systematic review and meta-analysis of global longitudinal strain and ejection fraction. Heart. 2014;100(21):1673–80.

4. Schelbert EB, Fridman Y, Wong TC, Abu Daya H, Piehler KM, Kadakkal A, et al. Temporal Relation Between Myocardial Fibrosis and Heart Failure With Preserved Ejection Fraction: Association With Baseline Disease Severity and Subsequent Outcome. JAMA Cardiol. 2017;2(9):995–1006.

5. Aro AL, Reinier K, Rusinaru C, Uy-Evanado A, Darouian N, Phan D, et al. Electrical risk score beyond the left ventricular ejection fraction: prediction of sudden cardiac death in the Oregon Sudden Unexpected Death Study and the Atherosclerosis Risk in Communities Study. Eur Heart J. 2017;38(40):3017–25.

6. Schelbert EB, Piehler KM, Zareba KM, Moon JC, Ugander M, Messroghli DR, et al. Myocardial Fibrosis Quantified by Extracellular Volume Is Associated With Subsequent Hospitalization for Heart Failure, Death, or Both Across the Spectrum of Ejection Fraction and Heart Failure Stage. J Am Heart Assoc. 2015;4(12).

7. Piehler KM, Wong TC, Puntil KS, Zareba KM, Lin K, Harris DM, et al. Free-breathing, motion-corrected late gadolinium enhancement is robust and extends risk stratification to vulnerable patients. Circ Cardiovasc Imaging. 2013;6(3):423–32.

8. Arheden H, Saeed M, Higgins CB, Gao DW, Bremerich J, Wyttenbach R, et al. Measurement of the distribution volume of gadopentetate dimeglumine at echo-planar MR imaging to quantify myocardial infarction: comparison with 99mTc-DTPA autoradiography in rats. Radiology. 1999;211(3):698–708.

9. Kors JA, van Herpen G, Sittig AC, van Bemmel JH. Reconstruction of the Frank vectorcardiogram from standard electrocardiographic leads: diagnostic comparison of different methods. Eur Heart J. 1990;11(12):1083–92.

10. Schlegel TT, Kulecz WB, Feiveson AH, Greco EC, DePalma JL, Starc V, et al. Accuracy of advanced versus strictly conventional 12-lead ECG for detection and screening of coronary artery disease, left ventricular hypertrophy and left ventricular systolic dysfunction. BMC Cardiovasc Disord. 2010;10:28.

11. Peduzzi P, Concato J, Feinstein AR, Holford TR. Importance of events per independent variable in proportional hazards regression analysis. II. Accuracy and precision of regression estimates. J Clin Epidemiol. 1995;48(12):1503–10.

12. Youden W. Index for rating diagnostic tests. Cancer. 1950;3(1):32–5.

13. Robin X, Turck N, Hainard A, Tiberti N, Lisacek F, Sanchez JC, et al. pROC: an open-source package for R and S+ to analyze and compare ROC curves. BMC Bioinformatics. 2011;12:77.

14. Oehler A, Feldman T, Henrikson CA, Tereshchenko LG. QRS-T angle: a review. Ann Noninvasive Electrocardiol. 2014;19(6):534–42.

15. Voulgari C, Pagoni S, Tesfaye S, Tentolouris N. The spatial QRS-T angle: implications in clinical practice. Curr Cardiol Rev. 2013;9(3):197–210.

16. Pavri BB, Hillis MB, Subacius H, Brumberg GE, Schaechter A, Levine JH, et al. Prognostic value and temporal behavior of the planar QRS-T angle in patients with nonischemic cardiomyopathy. Circulation. 2008;117(25):3181–6.

17. Bazett, HC. An analysis of the time-relations of electrocardiograms. Heart. 1920; 7:353.

18. Montanez A, Ruskin JN, Hebert PR, Lamas GA, Hennekens CH. Prolonged QTc interval and risks of total and cardiovascular mortality and sudden death in the general population: a review and qualitative overview of the prospective cohort studies. Arch Intern Med. 2004;164(9):943–8.

19. Gleeson S, Liao YW, Dugo C, Cave A, Zhou L, Ayar Z, et al. ECG-derived spatial QRS-T angle is associated with ICD implantation, mortality and heart failure admissions in patients with LV systolic dysfunction. PLoS One. 2017;12(3):e0171069.

20. Borleffs CJ, Scherptong RW, Man SC, van Welsenes GH, Bax JJ, van Erven L, et al. Predicting ventricular arrhythmias in patients with ischemic heart disease: clinical application of the ECG-derived QRS-T angle. Circ Arrhythm Electrophysiol. 2009;2(5):548–54.

21. Okin PM, Malik M, Hnatkova K, Lee ET, Galloway JM, Best LG, et al. Repolarization abnormality for prediction of all-cause and cardiovascular mortality in American Indians: the Strong Heart Study. J Cardiovasc Electrophysiol. 2005;16(9):945–51.

22. Broberg CS, Chugh SS, Conklin C, Sahn DJ, Jerosch-Herold M. Quantification of diffuse myocardial fibrosis and its association with myocardial dysfunction in congenital heart disease. Circ Cardiovasc Imaging. 2010;3(6):727–34.

23. Stanton T, Leano R, Marwick TH. Prediction of all-cause mortality from global longitudinal speckle strain: comparison with ejection fraction and wall motion scoring. Circ Cardiovasc Imaging. 2009;2(5):356–64.

24. Ng ACT, Prihadi EA, Antoni ML, Bertini M, Ewe SH, Ajmone Marsan N, et al. Left ventricular global longitudinal strain is predictive of all-cause mortality independent of aortic stenosis severity and ejection fraction. Eur Heart J Cardiovasc Imaging. 2018;19(8):859–67.

25. D’Agostino RB, Vasan RS, Pencina MJ, Wolf PA, Cobain M, Massaro JM, et al. General cardiovascular risk profile for use in primary care: the Framingham Heart Study. Circulation. 2008;117(6):743–53.

26. Guyon I, Elisseeff A. An introduction to variable and feature selection. Journal of machine learning research; 2003, 3(Mar). p. 1157–82.

27. Dastidar AG, Baritussio A, De Garate E, Drobni Z, Biglino G, Singhal P, et al. Prognostic Role of Cardiac MRI and Conventional Risk Factors in Myocardial Infarction With Nonobstructed Coronary Arteries. JACC Cardiovasc Imaging. 2019.

28. Bacharova L, Chen H, Estes EH, Mateasik A, Bluemke DA, Lima JA, et al. Determinants of discrepancies in detection and comparison of the prognostic significance of left ventricular hypertrophy by electrocardiogram and cardiac magnetic resonance imaging. Am J Cardiol. 2015;115(4):515–22.

29. Benjamin EJ, Wolf PA, D’Agostino RB, Silbershatz H, Kannel WB, Levy D. Impact of atrial fibrillation on the risk of death: the Framingham Heart Study. Circulation. 1998;98(10):946–52.

