## Supplementary material for "An electrocardiography score predicts heart failure hospitalization or death beyond that of cardiovascular magnetic resonance imaging": Detailed description of CMR methodology

**Supplemental material: Detailed description of CMR methodology**

CMR images were acquired using a 1.5 Tesla scanner (Magnetom Espree, Siemens Healthcare, Erlangen, Germany) and a 32-channel phased array cardiovascular coil. Examinations included standard breath held segmented cine imaging with steady-state free precession (SSFP) (1). LV mass, volumes and ejection fraction were measured from short-axis stacks of end-systolic and end-diastolic cine frames. LGE imaging was performed ten minutes after a 0.2 mmol/kg intravenous gadoteridol bolus (Prohance, Bracco Diagnostics, Princeton, NJ, USA), with a phase-sensitive inversion recovery pulse sequence to optimize LGE by rendering signal intensity proportional to T1 recovery. When patients could not breath hold, single-shot SSFP, and averaged phase-sensitive inversion recovery, motion corrected images were acquired (2). Typical acquisition parameters have previously been described (1).

*Quantification of the myocardial extracellular volume fraction*

An ECG-gated single-shot-modified Look Locker inversion recovery sequence (MOLLI) was used to acquire quantitative T1 maps. A native T1 map was acquired followed by a post-contrast T1 map after a gadolinium bolus injection. ECV was calculated in myocardium by delineating areas without LGE and calculated as: $ECV=\lambda\cdot(1-hematocrit)$, where λ = ΔR1myocardium/ΔR1bloodpool and ΔR1=1/T1_postcontrast_ - 1/T1_precontrast_ (3). Myocardial infarction and non-ischemic scar were defined as areas with LGE, and the middle third of the myocardium was traced to avoid partial volume effects. For blood ECV calculations, a circular region was traced in the middle of the blood pool to avoid partial volume effects by papillary muscles. The final ECV values were averaged from the basal and mid-ventricular short axis slices. Hematocrit measures were acquired on the day of CMR scanning. CMR data were analyzed using a commercial workstation (Leonardo, Siemens Healthcare, Erlangen, Germany).

*Quantification of global longitudinal strain*

GLS analysis was performed using semi-automated tissue feature tracking software (CVi42, Circle Cardiovascular Imaging Inc., Calgary, Canada). Epicardial and endocardial borders in the end-diastolic phase were manually traced in 2-, 3-, and 4-chamber views. Strain analyses tracings were inspected visually throughout the cardiac cycle, and manual changes were made when traces deviated from myocardial movement.

**References Supplemental material**

1.	Piehler KM, Wong TC, Puntil KS, Zareba KM, Lin K, Harris DM, et al. Free-breathing, motion-corrected late gadolinium enhancement is robust and extends risk stratification to vulnerable patients. Circ Cardiovasc Imaging. 2013;6(3):423-32.

2.	Kellman P, Larson AC, Hsu LY, Chung YC, Simonetti OP, McVeigh ER, et al. Motion-corrected free-breathing delayed enhancement imaging of myocardial infarction. Magn Reson Med. 2005;53(1):194-200.

3.	Arheden H, Saeed M, Higgins CB, Gao DW, Bremerich J, Wyttenbach R, et al. Measurement of the distribution volume of gadopentetate dimeglumine at echo-planar MR imaging to quantify myocardial infarction: comparison with 99mTc-DTPA autoradiography in rats. Radiology. 1999;211(3):698-708.
